# Emergence and transmission dynamics of the FY.4 Omicron variant in Kenya

**DOI:** 10.1101/2024.12.05.24318558

**Authors:** Sebastian Musundi, Mike J. Mwanga, Arnold W. Lambisia, John Mwita Morobe, Nickson Murunga, Edidah Moraa, Leonard Ndwiga, Robinson Cheruyoit, Jennifer Musyoki, Martin Mutunga, Laura M Guzman-Rincon, Charles Sande, Joseph Mwangangi, Philip Bejon, Lynette Isabella Ochola-Oyier, D James Nokes, Charles N. Agoti, Joyce Nyiro, George Githinji

## Abstract

The recombinant FY.4 SARS-CoV-2 variant was first reported in Kenya in March 2023 and was the dominant circulating variant between April and July 2023. The variant was characterised by two important mutations: Y451H in the receptor binding domain of the spike protein and P42L in open reading frame 3a. Using phylogenetics and phylodynamic approaches, we investigated the emergence and spread of the FY.4 in Kenya and the rest of the world. Our findings suggest FY.4 circulated early in Kenya before export to North America and Europe. Early circulation of FY.4 in Kenya was predominantly observed in the coastal part of the country and the estimated time to the most recent common ancestor suggests FY.4 circulated as early as December 2022. The collected genomic and epidemiological data show that the FY.4 variant led to a large local outbreak in Kenya and resulted in localised outbreaks in Europe, North America and Asia-pacific. These findings underscore the importance of sustained genomic surveillance especially in under sampled regions in deepening our understanding of the evolution and spread of SARS-CoV-2 variants.

## Introduction

Severe acute respiratory syndrome coronavirus 2 (SARS-CoV-2) emerged in Wuhan China in late 2019 (Lu et al., 2020; Zhu et al., 2020). Over the past five years, SARS-CoV-2 has evolved into several lineages, some of which are more transmissible than others and reduce effectiveness to public health interventions such as therapeutics, diagnostics and vaccines (Centre for Disease Control and Prevention, 2021). The World Health Organization (WHO) designated these lineages as variant of concern (VOC) (https://www.who.int/activities/tracking-SARS-CoV-2-variants). The VOCs include Alpha (B.1.17) (Hill et al., 2022), Beta (B.1.35) (Tegally et al., 2021), Gamma (P.1) (Faria et al., 2021), Delta (B.1.617.2) (Cherian et al., 2021) and Omicron(Viana et al., 2022). The Omicron variant was first reported on 24th November 2021 in South Africa and Botswana and carried over 30 mutations in the spike protein compared to the Wuhan-Hu-1 strain (Tegally et al., 2022; Viana et al., 2022). The presence of these mutations increased transmissibility and reduced neutralization by anti-SARS-CoV-2 antibodies (Planas et al., 2022). While the origin of the Omicron remains debatable, possible hypotheses explaining its emergence include extreme under-sampling resulting in undetected lineages, zoonotic spillover from animals that transmit SARS-CoV-2 without an intermediate host like the white-tailed deer and farmed mink and persistence of the virus in chronically infected individuals (Markov et al., 2023).

The Omicron variant was initially classified into three lineages BA.1, BA.2 and BA.3 before the emergence of BA.4 and BA.5 (Tegally et al., 2022). Unlike previous waves of VOCs which were driven by few circulating variants (Li et al., 2021), the emergence of Omicron led to convergent evolution and diversification (Andreis et al., 2023; Ito et al., 2023), resulting in multiple co-circulating lineages and an increased probability of recombination events. In mid-August 2022, the XBB recombinant was reported in India, Singapore and other parts of Asia (Wang et al., 2023). XBB emerged from two BA.2 omicron variants BM.1.1.1 (BA.2.75.3.1.1.1) and BJ.1 (BA.2.10.1.1) with breakpoints located in the receptor-binding domain of spike protein (Tamura et al., 2023). XBB showed increased immune resistance, fusogenicity and binding affinity to angiotensin-converting enzyme 2(ACE2), the receptor mediating viral infection compared to the parental strains (Tamura et al., 2023). Since then, multiple XBB variants such as XBB.1.5 (Parums, 2023), and XBB.1.16 (Looi, 2023) emerged and spread across the globe.

The FY.4 (XBB.1.22.1.4) omicron variant became the dominant lineage in Kenya between March to July 2023 in Kenya following its initial detection on 10^th^ March 2023 (Mwanga et al., 2023). Globally, the first FY.4 sample outside Kenya was reported first on 3^rd^ April 2023 in Germany (Khare et al., 2021). In this work, we aimed to understand the origin and transmission of the FY.4 Omicron sub-variant using genomic data. FY.4 circulated as early as December 2022 and was observed in Kenya first before being exported to other regions of the world. These findings suggest FY.4 potentially originated from Kenya and underscores the importance of genomic surveillance in tracking SARS-CoV-2 variants.

## Methods

### Data sources

SARS-CoV-2 FY.4 samples (n=73) used for analysis in this study were generated at Kenya Medical Research Institute Wellcome Trust Research Programme (KWTRP) as previously described (Mwanga et al., 2023). An additional (n=139) FY.4 genomes sequenced within Kenya and across the globe were retrieved from Global Initiative for Sharing all Influenza Data (GISAID). Consensus genomes for FY.4 lineages and XBB were retrieved from (https://github.com/corneliusroemer/pango-sequences) and aligned using MAFFT. Amino acid mutations relative to the Wuhan-HU-1 and XBB and were plotted using SNIPit (O’Toole et al., 2024).

## Time-scaled analysis of the FY.4 variant

We compiled a dataset of circulating SARS-CoV-2 sequences in Kenya between October 2022 and October 2023 (n=1,078). Out of these 73 genomes were sequenced at KWTRP and an additional (n=139) sequenced locally in Kenya, making a total of 212 Kenyan FY.4 sequences. We analysed this dataset against a background of FY.4 global sequences retrieved from GISAID (n=755). The global dataset from where the sequences were obtained are summarized in Supplementary Table 1. Sequences with less than 70% genome coverage (n=1), incomplete collection dates (n=11), and whose lineage were misclassified based on NextClade version 3.8.2 (n=5) were excluded from further analysis. The remaining sequences were aligned against the SARS-CoV-2 (XBB) reference (a Wuhan-Hu-1/2019 reference (Accession number MN908947.3) containing XBB single nucleotide polymorphisms) using MAFFT version 6.240 (Katoh & Standley, 2013). The alignment was visualized using AliView version 1.28 (Larsson, 2014) and the 5’ and 3’ ends containing the untranslated regions were manually trimmed. A maximum likelihood (ML) tree was generated using IQ-TREE version 2.3.3 with 1000 bootstrap values using the General Time Reversible (GTR) model (Nguyen et al., 2015). The maximum likelihood tree was visualized using the R package “ggtree” and low-quality sequences (n=16) dropped from the tree using the R package “ape.” A time-resolved maximum likelihood tree was then generated using Treetime (Sagulenko et al., 2018).

## Phylogeography analysis

The presence of a molecular signal was evaluated using TempEst version 1.5.3 (Rambaut et al., 2016) prior to Bayesian analysis using BEAST (Drummond & Rambaut, 2007). To identify the clades circulating within Kenya, a discrete phylogeography approach utilizing two locations; “Kenya” and “Others” was run in BEAST 1.10.4. This approach utilized the time-resolved phylogeny as the starting tree to reduce computational time needed to generate transmission information(Bollen et al., 2021). Bayesian inference was run using BEAST v1.10.4 for 6 x 10^5^ Monte Carlo Markov Chain (MCMC) and sampling every 1000 step. Log files were checked for convergence and mixing using Tracer v1.7.1 (Rambaut et al., 2018). After discarding 10% as burn-in, a maximum clade credibility tree (MCC) generated using TreeAnnotator version 1.10.4 (Suchard et al., 2018). The overall number of transitions between the two locations was counted in BEAST package Babel utilizing sampled trees.

To explore the local transmission of the FY.4 variant within Kenya, the relaxed random walk (RRW) diffusion model which allows for dispersal velocity in the tree to vary but remain the same in the branches was inferred using the longitude and latitudes of sample locations as continuous traits (Lemey et al., 2010; Pybus et al., 2012). Since the RRW model does not allow samples to have the same geographical coordinates, we selected the centroid location of each administrative point and added random jitter of 0.05 to each tip making each location distinct. A MCMC chain was run in duplicate for 500 million iterations sampled after every 50,000 steps with its mixing properties checked in Tracer v.1.71 to ensure the effective sample size (ESS) is >200. The R package “seraphim” used to extract spatiotemporal information embedded in the posterior, maximum clade credibility trees and visualize the dispersal of FY.4 within Kenyan counties (Dellicour et al., 2016).

Next, we carried out a discrete phylogeographic analysis to infer the global origin and movement of FY.4 across different regions across the globe. We aggregated the country of sampling into four regions including Kenya (n=206), North America (n=494), Europe (n=140) and Asia-Pacific (Asia and Oceania) (n=91). One sample from South America and two samples from Uganda were excluded to minimize biased posterior distributions due to unequal sample size. In addition, we analyzed samples collected from March 2023 considering the first case was reported on 10^th^ March 2023. By focusing on this timeline, we aimed to capture the spread of FY.4 from the time it was initially identified providing a clear evolutionary trajectory of the geographic spread of FY.4 while in circulation. During this study, three additional FY.4 sequences were recently deposited in GISAID from Nakuru, Kenya with collection dates ranging between 11^th^ -14^th^ November 2022. Although these sequences predate the first reported case suggesting earlier circulation, we excluded these samples due to insufficient information from very few samples collected in November 2022 and absence of samples collected between November 2022 and March 2023. Since a large number of sequences originated from the USA, down sampling was carried out based on time, pangolin lineage and region resulting in a total of 511 sequences distributed as follows: Kenya (n=94), North America (n=233), Europe (n=107) and Asia-Pacific (n=77). Ancestral locations reconstruction of the FY.4 variant and asymmetric viral exchanges between regions was estimated utilizing a non-continuous time Markov chain model. The Bayesian stochastic search variable selection was used to infer non-zero migration rates and identify the best-supported transitions rates using a bayes factor test. In addition, Markov jump counting was used to estimate the number of transitions across the different regions. The time-scaled phylogeny was created using the HKY substitution model with a gamma distributed rate for invariant sites, an uncorrelated relaxed molecular clock with a log normal prior and Bayesian skygrid with 52 change points over a period of one year. The analysis was run in duplicate on BEAST v1.10.4 using 200 million MCMC steps with sampling after every 20000 steps. Mixing and convergence properties were assessed using Tracer v.1.71 ensuring the ESS was >200. After discarding the 10% burn-in, the MCC tree was constructed using TreeAnnotator version 1.10 and the number of transitions/Markov jumps estimated using TreeMarkovJumpHistoryAnalyzer (Lemey et al., 2021).

## Results

### FY.4 circulated in Kenya before global expansion and underwent rapid evolution over a short period of time

FY.4 variant became the dominant circulating lineage in Kenyan between March and July 2023 before eventual replacement with other Omicron subvariants (Figure 1A). The period coincides with the high positivity rate observed from samples collected from health facilities within the Kilifi Health Demographic Surveillance System (KHDSS) (Supplementary Figure 1). On the global scale, FY.4 was first detected in Africa before its eventual detection in other continents across the globe (Figure 1B). In Africa, FY.4 circulation peaked between April and May 2023 while in North America and Europe the peak was observed in July 2023 (Figure 1B). In Africa, sublineage FY.4.1 was the dominant circulating lineage and co-circulated with its expanded lineage FY.4.1.2. North America initially saw FY.4.1 and FY.4.2 in April 2023 followed by FY.4.1.2 before the co-circulation of FY.4.2 and FY.4.1.1 in the later stages. FY.4.1 and FY.4.1.2 were the main circulating lineages in Europe from April 2023 and while the same lineages were initially detected in Asia from May 2023, FY.4.2 became the main circulating lineage thereafter from June to September 2023 (Figure1B).

**Figure 1.**
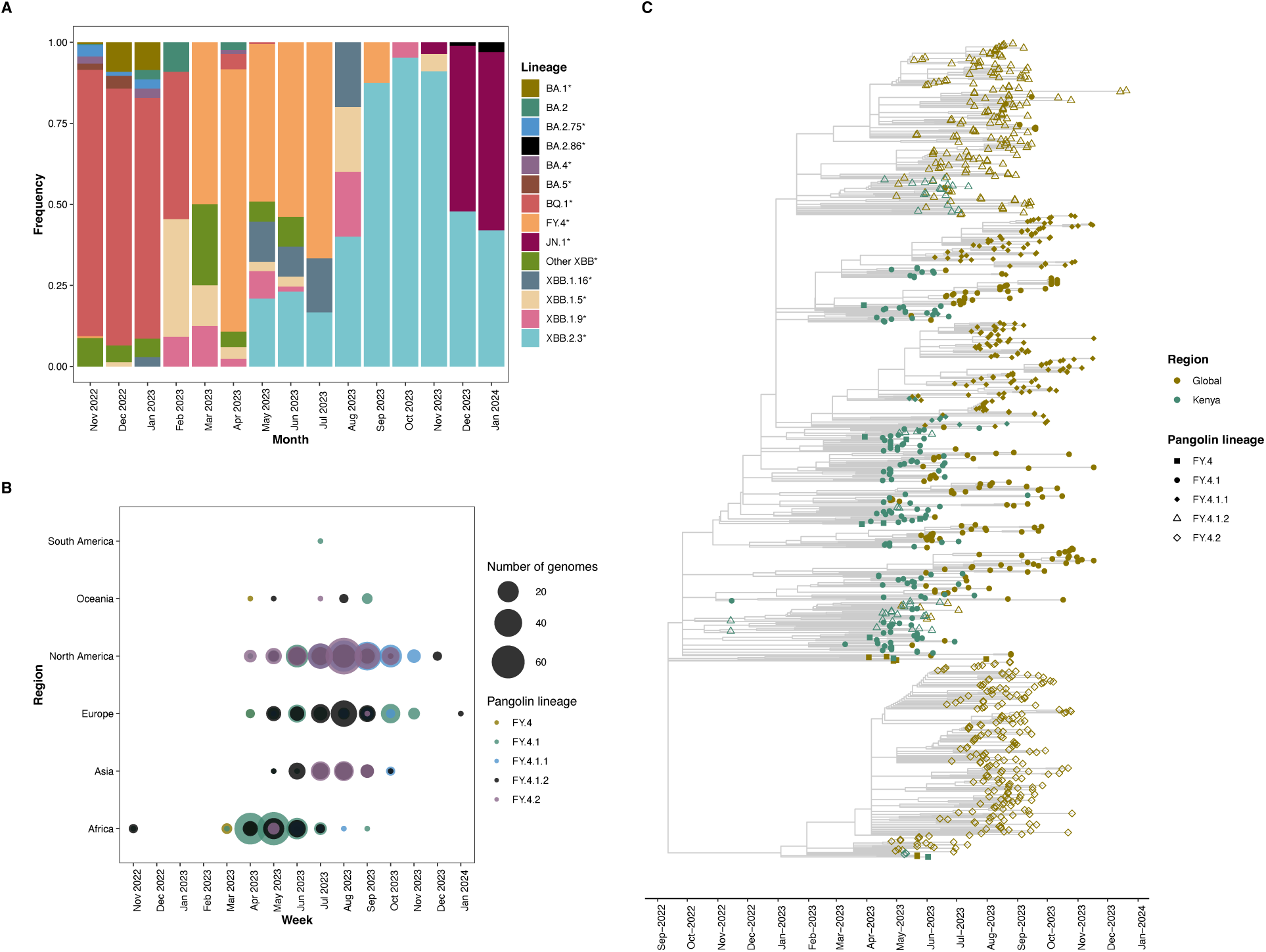
Circulation of the FY.4 omicron subvariant. A) Proportion of SARS-CoV-2 lineages circulating in Kenya from September 2022 to January 2024. FY.4 circulated in Kenya predominantly from March to September 2023. B) Global distribution of circulating FY.4 variants from November 2022 to January 2024. The individual colours represent the FY.4 sub-lineages while the size of the circle denotes the number of genomes deposited in GISAID per month. C) Time-resolved phylogenetic tree showing the temporal clustering of FY.4 between Kenya and global sequences. The colours represent the location the sample was collected from, and the shape represent the FY.4 pangolin sub-lineages.

Phylogenetic analysis of the circulating strains in Kenya (n=1078) between October 2022 and October 2023, showed that FY.4 was closely related to other XBB variants and more specifically XBB.1.16 and XBB.1.9 (Supplementary Figure 2). As expected, FY.4 was also present in the same clade as BA.2 and BA.2.75 which has previously been proposed as the main parental lineages for the emergence of the recombinant XBB. In addition to the two lineage-defining mutations-Y451H in the spike protein and P42L in ORF3a- expanded sub-lineages exhibited notable mutations. FY.4.1 contained S494P in the receptor binding domain of spike glycoprotein while FY.4.1.1 included S704L in subdomain 2. The FY.4.1.2 sub-lineage was characterized by S2926F in ORF1a and FY.4.2 displayed mutations Y2171F in ORF1a and V2287I in ORF1b.

A time-resolved tree ML tree containing Kenyan (n=207) FY.4 and global sequences (n=738) revealed a short period between the expansion of FY.4.1 to FY.4.1.1 and FY.4.1.2 suggesting a rapid diversification process (Figure 1C). Following its expansion, FY.4.1 persisted for 36 weeks in circulation together with its derived lineages. In contrast, FY.4.2, which also emerged from FY.4, did not expand to additional lineages, likely reflecting the different selective pressures acting on individual FY.4 lineages. Analysis of the temporal spread revealed FY4 circulated early in Kenya compared to other regions of the globe (Figure 1C). Additionally, Kenyan FY.4 sequences displayed significant genetic diversity by mapping to multiple lineages and formed clusters providing evidence for local transmission ahead of introductions to other regions (Figure 1C).

## The FY.4 variant originated in Kenya

The presence of a temporal signal was confirmed by the linear relationship between genetic divergence and sampling dates using TempEst version 1.5.3 (R^2^=0.326) (Figure 2A). Subsequently, we carried out a discrete phylogeographic analysis to identify the potential origin and the number of import and export events to Kenya using the FY.4 variant using genomic data. Based on this, we observed no evidence of independent introductions of FY.4 in Kenya (Figure 2B) based on the 95% highest posterior density (HPD) ([0-1]) However, we identified at least 75 transmission events from Kenya to the rest of the globe (95% HPD interval = [73-79]). Local transmission dynamics in Kenya, comprised of over 485 transition events between counties (95% HPD interval= [480-489]) (Figure 2B). Evidence of spread is supported by export and imports events between the counties. Global spread was observed with support for multiple export events (1,304 transition events (95% HPD interval = [1298-1312]) across the globe showing FY.4 arrived and transmitted across multiple regions.

**Figure 2.**
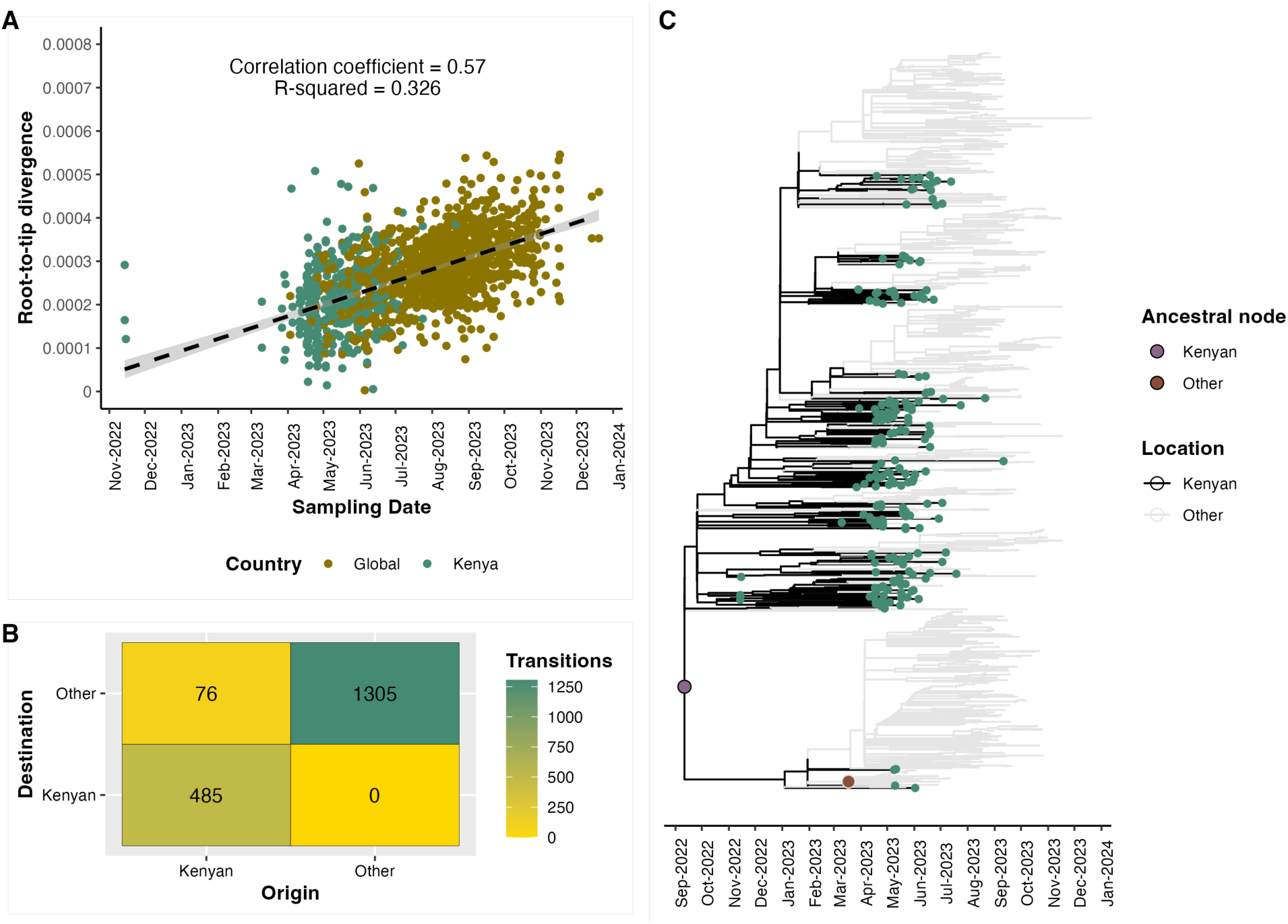
Preliminary phylogeography discrete trait analysis. A) Root-to-tip regression from TempEst analysis showing the presence of a temporal signal (R^2^ =0.326, correlation coefficient = 0.57). The FY.4 sequences are colored according to the locations (Kenya or Global). The regression line represents the estimate mean evolutionary rate with error buffers in grey showing the 90% confidence intervals. B). The number of transitions was derived from the MCC tree by counting changes between “Kenyan” and “Other” locations. Transitions were counted when the location of the internal node changed from “Kenya” to “Others” or vice versa or when maintained in the same position. C) Preliminary discrete trait analysis identified two ancestral clades associated with spread of FY.4 using a fixed time-resolved tree. The green tips represent individual Kenya samples, black the most probable location for Kenyan sequences and background grey the most probable location for global sequences. Two ancestral nodes are filled by the plum and salmon colours respectively. The “Other” ancestral node occurs primarily on the FY.4.2 sub- lineage which was not largely observed in Kenya but was predominant in North America.

Ancestral reconstruction analysis revealed the presence of two most common recent ancestors. The first node infers Kenyan ancestry and included all Kenyan sequences alongside global sequences forming the largest clade with 915 sequences (Figure 2C). The genetic diversity of the Kenyan sequences in this node supports strong local transmission within Kenya while the presence of both Kenyan and global sequences in the same node suggests possible worldwide dissemination of the FY.4 variant. In contrast, the second ancestral node was associated with fewer sequences (n=19) and likely represented a more localized transmission route. Notably, the second ancestral node did not include Kenyan samples which might suggest limited or no transmission to or from Kenya or a pathway that completely bypassed Kenya (Figure 2C).

We extracted the ancestral clade that inferred Kenyan ancestry and examined virus dispersal within Kenya. The subsequent continuous phylogeography of patterns of dispersal within Kenya supported the evidence that FY.4 was the dominant circulating lineage in coastal region from March 2023. By May-June 2023, most cases were observed centrally in the capital city (Figure 3).

**Figure 3.**
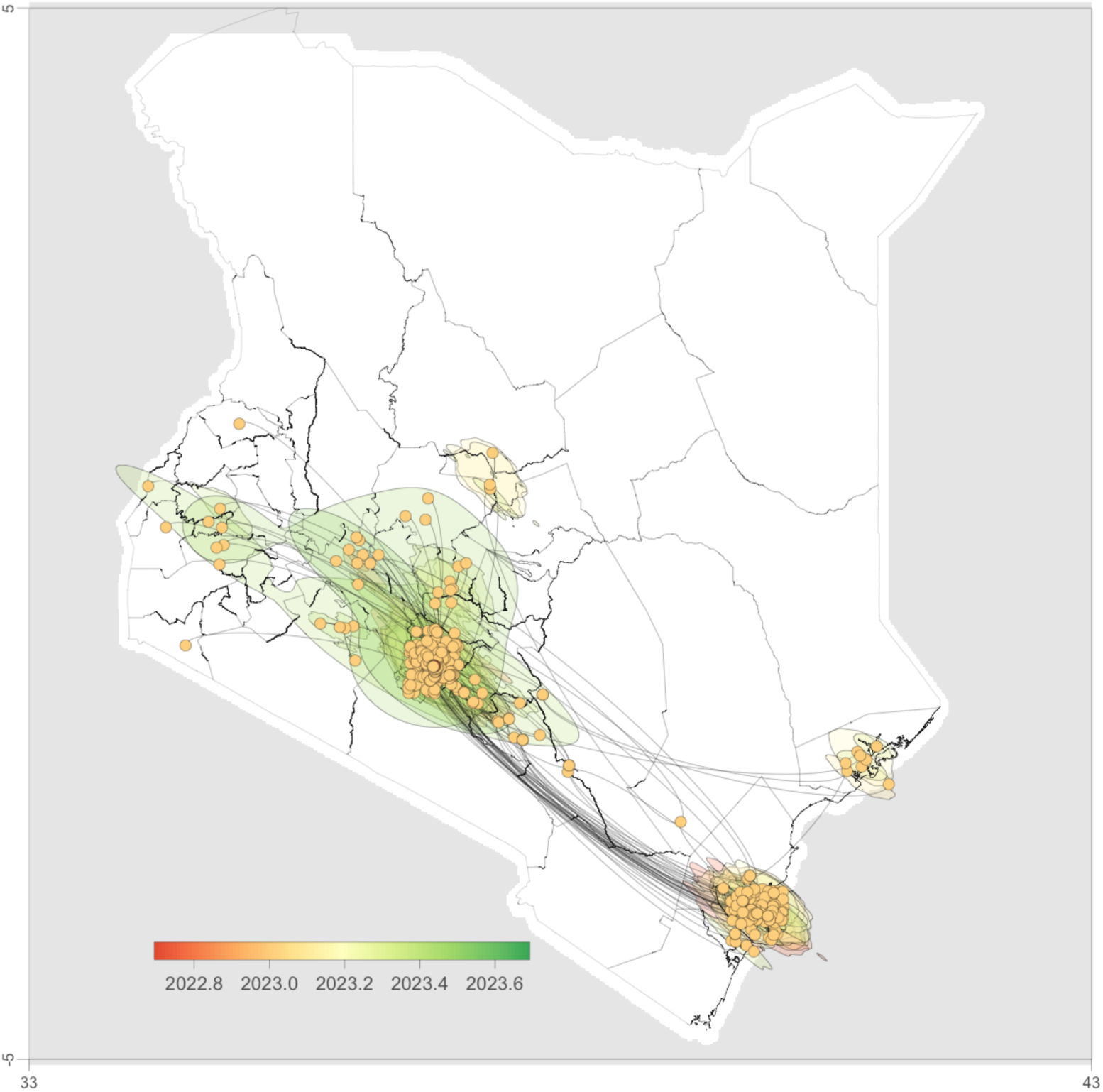
Dispersal of FY.4 across Kenya over time based on 1000 subsampled trees from a continuous phylogeographic posterior distribution. The nodes of the MCC tree are color-coded based on the time of occurrence. The 80% HPD regions are displayed in successive layers with the colors reflecting corresponding time periods for virus spread.

Most peripheral samples also directly connected to either the coastal region or centrally in the capital city which were the predominant sampling sites. This pattern likely suggests a limited geographical range of sampling in Kenya, potentially leading to an underestimate of the circulation of FY.4 in other parts of Kenya.

Phylogeographic reconstruction implied that FY.4 may have emerged in Kenya in early January (mean tMRCA 3^rd^ January, 95% HPD 2^nd^ December 2022- 1^st^ February 2023) (Figure 4A). Thereafter, FY.4 was first exported to Europe then to North America in week 7 and 9 respectively and later exported to Asia-Pacific around week 13 (Figure 4B). Out of the 60 export events from Kenya, most were to North America (n=32), followed by Europe (n=19) and lastly Asia-Pacific (n=9) (Figure 4B). There were no inferred introductions from the globe into to Kenya during this period. An exponential increase in the effective population size was observed between January and April 2023 followed by a minor decline and another slight exponential increase from August to September before a decline to January 2024 (Figure 4C).

**Figure 4.**
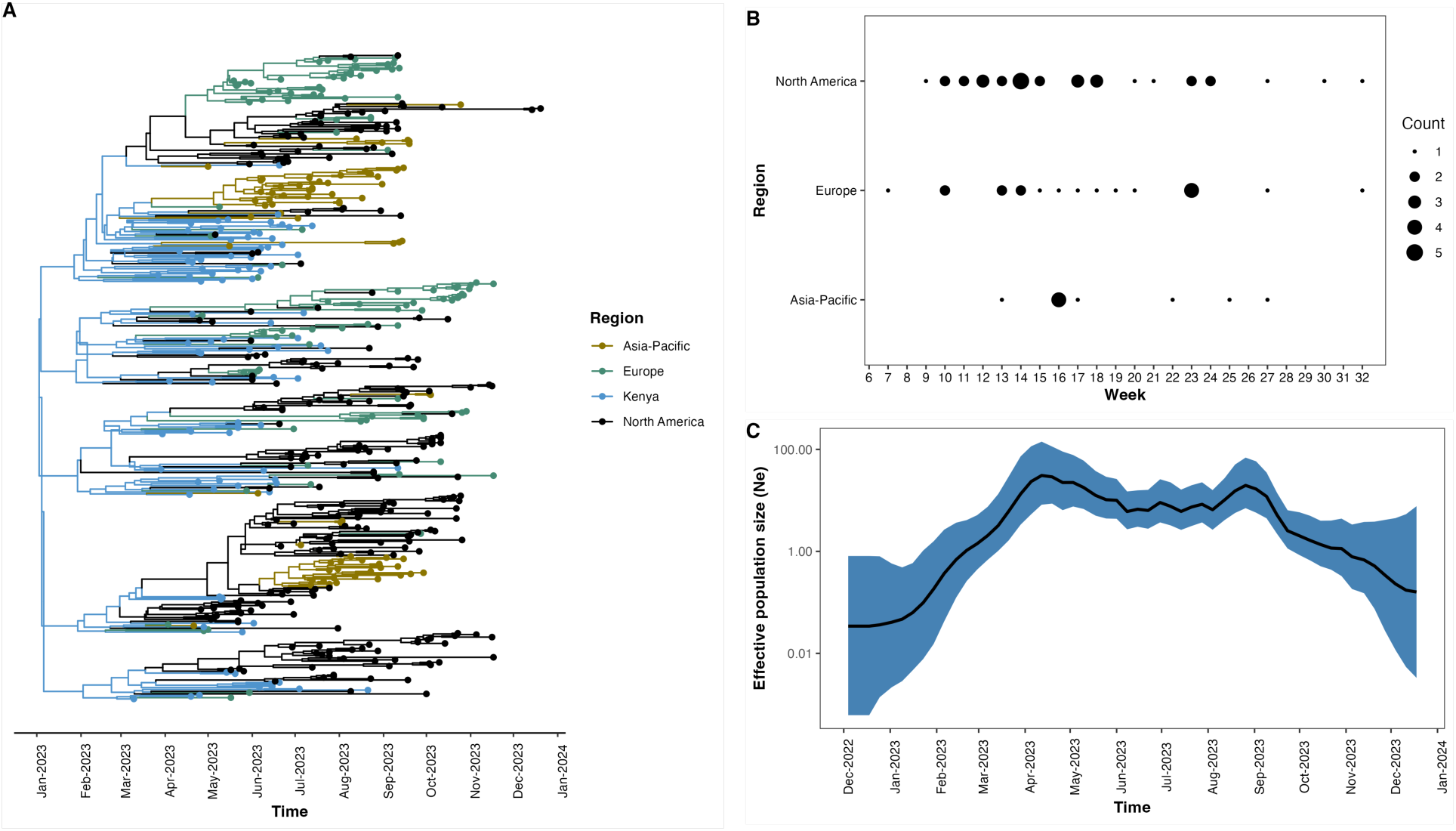
Bayesian phylogeographic reconstruction of FY.4. A) Time-resolved maximum credibility clade tree with branches coloured by inferred geographic location. B) A summary of the number of Markov Jumps from Kenya to other regions stratified by epidemiological weeks. C) A Bayesian Skyline plot describing the inferred change in the effective population size of FY.4 infections over time.

## Discussion

The FY.4 variant increased in circulation in Kenya between March and July 2023 accounting for increased cases and hospitalization (Mwanga et al., 2023). This variant was also observed in other regions across the globe (Figure 1C). Given the early isolation of FY.4 and increased transmission intensity in Kenya, we applied genomic and epidemiological data to investigate the emergence and transmission dynamics of FY.4 variant from sequenced samples in Kenya in addition to those collected from the globe.

The FY.4 lineage of SARS-COV-2 is characterized by two notable mutations namely the Y451H in the receptor binding domain (RBD) of the spike protein, whose functional implication is unclear and the P42L in the ORF3a, with potential for contribution to the loss of recognition of T-cell epitope (de Silva et al., 2021). Additionally, the S494P present in the RBD of FY.4.1 enhances binding affinity to ACE2 receptor increasing transmissibility (Chakraborty, 2021) and potentially contributing to the increased number of observed cases. Neutralization assays against multiple circulating Omicron variants including FY.4 in Kenya between March 2023-March 2024 have provided evidence for decline in naturally acquired and vaccine-mediated antibody responses implying that the Kenyan population was still susceptible to infections caused by emerging Omicron subvariants. (Lugano et al., unpub. data doi.org/10.1101/2024.06.26.24309525). Here, we used an established health facilities surveillance platform (Nyiro et al., 2018), and combined this with further local and global genomic data to determine the origin and describe the transmission dynamics of this variant.

The phylogenetic and phylogeographic reconstruction suggests that Kenya was the potential origin of the FY.4 variant (Figure 2B). Phylogenetic and phylogenomic analysis provides evidence for multiple exportation events from Kenya, primarily to North America and Europe (Figure 3B) which occurred between March and July 2023. Previously we have observed multiple introductions of ancestral strains (Githinji et al., 2021) and variants of concern (Agoti et al., 2022; Githinji et al., unpub. data doi.org/10.1101/2022.10.26.22281446). The Bayesian skyline plot showed that the effective population size corresponded with increase in the number of cases and potentially indicate missed cases during the outbreak period. A peak in the effective population size was observe between August and September 2023 suggesting a surge in cases outside of Kenya and this coincides with a rise in the FY.4 genome sequences deposited from North America and Europe (Figure 3C). This suggests ongoing transmission events between Kenya and the rest of the world following the initial emergence of FY.4.

A limitation with this study is that the true number of FY.4 derived COVID-19 cases in Kenya, African and South America is likely underestimated given the limited number of tests at the period, under-surveillance and the limited genomic surveillance in the country. The biased sampling is underscored by clustered outbreaks in areas with higher sampling rates.

## Conclusion

Genomic surveillance is essential in identifying emerging variants of SARS-CoV-2. In this study, the use of phylogenetics and phylodynamic approaches provided insights to the possible origin and dispersal patterns of the FY.4 variant The study reinforces the need for increased sequencing capacity especially in under-sampled geographic regions especially in studying the evolution of SARS-CoV-2.

## Data Availability

The raw data files and consensus genome sequences obtained in this study were submitted to both GISAID and GenBank databases and the accession numbers prepared for deposition in Harvard DataVerse (https://doi.org/10.7910/DVN/VPWUXN). For more detailed information beyond the metadata used in the paper, there is a process of managed access requiring submission of a request form for consideration by our Data Governance Committee.

## Data Availability

All data produced in the present work are contained in the manuscript

## Acknowledgments

We thank the members of our field study team involved in collecting the samples, the members of the KWTRP COVID-19 testing team, PEO research group members that undertook real-time RT-PCR processing. This work was support by multiple funding sources that included the New Variant Assessment Platform (NVAP), Wellcome grants (220985/Z/20/Z and 226002/A/22/Z). The Rockerfeller Foundation subaward (OXFFDG01) and the Department of Health and Social Care grant (project references 17/63/82 and 16/136/33). The views expressed in this publication are those of the author (s) and not necessarily those of the Department of Health and Social Care, Foreign Commonwealth and Development Office, Wellcome Trust or the UK government.

This manuscript was published with permission from the Director General

## Author’s contributions

The project was conceived and designed by GG; Project supervision and funding was obtained by GG, CNA, LI, JN, DJN; Laboratory processing of the samples was conducted by MM, AWL, JM, EM, LK, RC, JM, MM, LG; Data management and analysis was carried out by NM,SM,LN. SM,GG wrote the initial draft and GG, CNA, PB, reviewed the manuscript and produced the final draft. All authors contributed to and reviewed the final draft.

