## Supplementary material for "Emergence and transmission dynamics of the FY.4 Omicron variant in Kenya"

**Supplementary Figure 1. Number of samples collected, tested and positivity rate in KHDSS in the first 26 weeks of 2023.** The line graph represents the number of samples collected each week while the bar graph shows the positivity rate within the same period. A total of 125 out of 1934 (0.06%) SARS-CoV-2 samples collected within KHDSS between January to July were positive for SARS-CoV-2 with the positivity rate peaking in April.

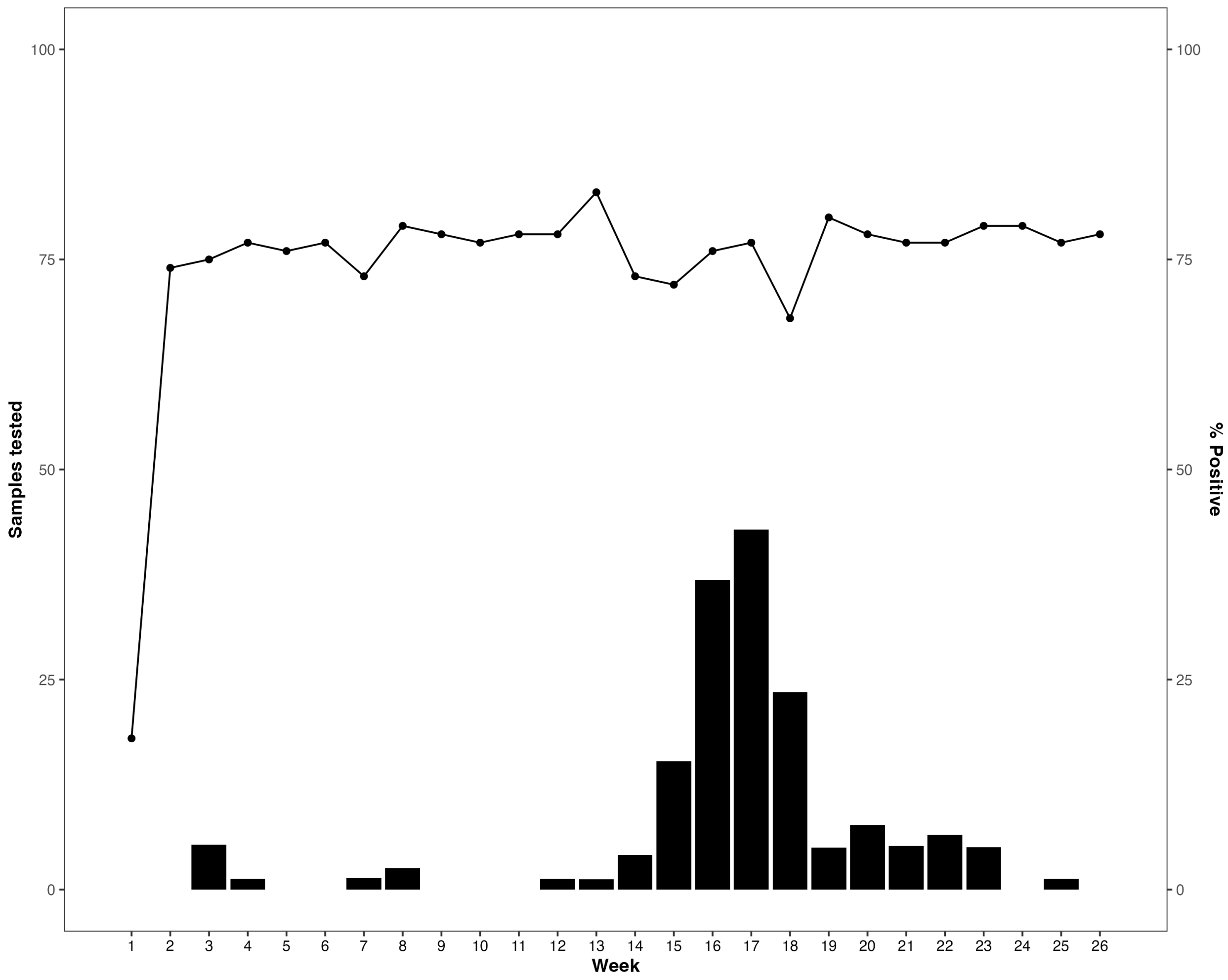

**Supplementary Figure 2. Time-calibrated tree showing the circulation of SARS-CoV-2 in Kenya between September 2022 to January 2024 using data deposited in GISAID**. Tips are colored based on the circulating lineages. FY.4 found in the same clade clustering with other XBB variants specifically XBB.1.9 and XBB.1.16. The parental lineages to XBB, BA.2 and BA.2.75 are also found in the same clade as FY.4.

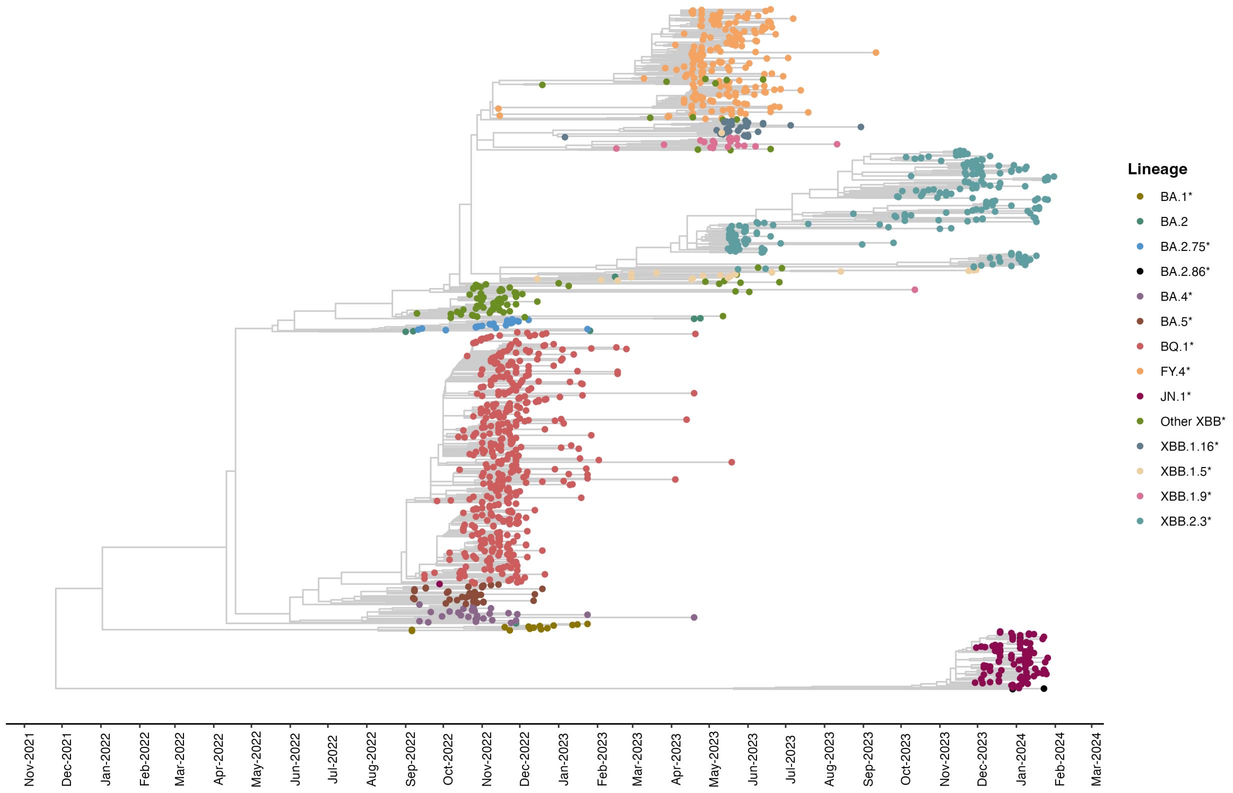

**Supplementary Table 1. Country of origin and number of FY.4 sequences retrieved from GISAID**.

| **Country** | **Number of samples (n=755)** |
|---|---|
| Australia | 8 |
| Austria | 3 |
| Belgium | 1 |
| Brazil | 1 |
| Canada | 43 |
| China | 5 |
| Croatia | 3 |
| Denmark | 2 |
| France | 6 |
| Germany | 4 |
| Greece | 1 |
| India | 1 |
| Indonesia | 1 |
| Ireland | 10 |
| Israel | 2 |
| Italy | 3 |
| Japan | 39 |
| Netherlands | 1 |
| Portugal | 1 |
| Puerto Rico | 1 |
| Romania | 3 |
| Saudi Arabia | 1 |
| Slovenia | 3 |
| South Korea | 36 |
| Spain | 2 |
| Sweden | 19 |
| Switzerland | 3 |
| Uganda | 2 |
| United Kingdom | 80 |
| US | 469 |
